# Experiences of frontline physiotherapists during the COVID-19 pandemic at two tertiary hospitals in Southern Malawi: A phenomenological qualitative study

**DOI:** 10.64898/2026.08.07.26359182

**Authors:** Allan Phiri, Emmanuel Chinula, Carolyn Jangwa, Ephraim Khosa, Nesto Tarimo, Fanuel Meckson Bickton

## Abstract

Frontline healthcare workers (HCWs) during the COVID-19 pandemic in Malawi included physiotherapists. This study explored the experiences of those physiotherapists to help prepare workforce support plans in the future in the advent of a new disease outbreak. This phenomenological qualitative study was conducted between 2 May and 15 June 2024 at two tertiary hospitals in Southern Malawi namely, Queen Elizabeth Central Hospital (QECH) and Zomba Central Hospital (ZCH). Participants were purposely sampled and included physiotherapists who had been involved in caring for patients with COVID-19 at the two hospitals. Data was collected from 11 participants (9 from QECH and 3 from ZCH) using physical in-depth interviews. The recorded interviews were transcribed verbatim and transcripts analyzed using a deductive thematic approach. Themes were broadly categorized into positive and negative experiences. Among positive experiences, participants reported that physiotherapy interventions facilitated quick recovery of patients. In some instances where oxygen cylinders were not enough or had run out of oxygen, physiotherapy interventions were lifesaving. Additionally, the COVID-19 pandemic raised awareness of physiotherapy’s role in COVID-19 management and resulted in permanent employment for several physiotherapists. Under the theme of negative experiences, participants faced challenges with team recognition, communication, staff shortages, inadequate equipment, and no local physiotherapy guidelines. The findings suggest Malawi’s healthcare system needs better pandemic preparedness and stronger interdisciplinary care.

## Introduction

Malawi is one of the countries which were affected by the Coronavirus 2019 (COVID-19) pandemic. As of 26 July 2025, there were 89,564 confirmed cases of COVID-19 and 2,687 COVID-19-related deaths reported in the country since the pandemic began (1). In response to the increasing number of hospitalized patients, the Malawi government in addition to already employed healthcare workers (HCWs) recruited additional ones including physiotherapists to alleviate the shortage of staff. Also, the makeshift facilities were built to accommodate the increased number of hospitalizations and effectively respond to the pandemic (2). To understand the HCW support needs during the COVID-19 crisis, several qualitative studies have been conducted in different countries. However, such studies are mostly limited to high-income countries; few have been conducted in low- and middle-income countries (3). Moreover, these studies have limited participant diversity, predominantly focusing on nurses and doctors compared to HCWs of other cadres such as physiotherapists (3–5).

To address the last research gap, a qualitative literature review by van der Westhuizen and Killingback (6) focused specifically on the experiences of physiotherapists who had worked directly with patients with COVID-19 during the COVID-19 pandemic. This was to understand whether there was a nuanced perspective of physiotherapists regarding their experiences during the COVID-19 pandemic (6). However, only six articles were eligible and included in the review, an indication of the limited number of studies available in this area. Of the six articles, only two reported studies from the continent of Africa, namely Hassem et al in South Africa (7) and Igwesi-Chidobe et al in Nigeria (8). This small number of African studies limits the transferability of the results to other African countries where physiotherapists may have their own stories to tell (9).

In Malawi, a few qualitative studies have been conducted to explore HCWs’ experiences during the COVID-19 pandemic. One such study was conducted among 25 HCWs at Kamuzu Central Hospital by Gundo et al (10). However, only 2 of the 25 participants were physiotherapists. In addition, the experiences of these two physiotherapists were not clearly reported as the authors did not include the HCW cadre of a participant alongside their quote. The other studies (9,11,12) did not include physiotherapists at all.

Hence, this undergraduate study was conducted to explore the experiences of physiotherapists who were involved in the management of patients with COVID-19 at two tertiary hospitals in southern Malawi. These experiences can inform the development of interventions for supporting physiotherapists in managing COVID-19 and future pandemics in Malawi, a key to strengthening health systems resilience (3).

## Methods

### Study type and design

This cross-sectional phenomenological qualitative study was conducted between 15 May 2024 and 15 June 2024. This study focused on understanding unique lived experiences of physiotherapists who were involved in management of patients with COVID-19, by exploring the meaning of the phenomenon and examining their experience through the descriptions provided by participants themselves (13). Researchers were meeting the participants only at a single point in time and the information collected was based on the participants’ experience.

### Study setting

The study was conducted at two tertiary hospitals in the southern region of Malawi which are Queen Elizabeth Central Hospital (QECH) and Zomba Central Hospital (ZCH). These two hospitals are amongst the four referral hospitals in the country, which offer comprehensive services which include drug administration, oxygen administration, feeding, continuous monitoring, mechanical ventilation in isolated cases and mental health services such as counselling. Critically ill COVID-19 patients from the southern region were being referred to these hospitals from different community health centers and some centers that were designed specifically for COVID-19 patients (12).

### Sampling method and sample size

Participants were recruited through non probabilistic purposive sampling techniques. The study included eleven qualified physiotherapists, of which eight were from Queen Elizabeth Central Hospital and three from Zomba Central Hospital. The physiotherapists who were actively involved in providing direct care for patients with COVID-19 during the first and second outbreak were initially eligible. This sample size was determined by data saturation, which was achieved at participant number nine, but the researchers continued to interview two more participants which brought the total number of participants to eleven.

### Ethics approval

Ethics approval was obtained from the Kamuzu University Research Ethics Committee, College of Medicine Research and Ethics Committee (CoMREC). Data collection was started after approval of the proposal by COMREC (Number P.10/23-0369). Before participants taking part in the study, they were clearly told about the purpose of the study, the benefits and the risks associated with taking part in the study. Identities of the participants were kept confidential by using codes. Hard copies of the participants were kept in lockers with keys. Soft copies were encrypted using passwords. This allowed only investigators and supervisors to have access to the documents and avoid any inconveniences which may have arisen due to unauthorized access. Participants voluntarily signed consent forms and had the right to withdraw from the research anytime they wanted. This is according to the declaration of Helsinki.

### Data collection

The potential participants were approached by the researchers and were provided with written information about the study. The health workers who expressed willingness to participate in the study provided written consent. The interviews were conducted at a convenient time and place for the participants within the hospital. They were conducted in English, and a semi-structured interview guide was used. The content of the interview guide covered the following areas: the experiences of the physiotherapist regarding working in multidisciplinary, exploring perspectives of the physiotherapists regarding the training provided to front-line healthcare workers in the of management of COVID-19 and exploring the views of physiotherapists regarding the performance of healthcare system in the management of patients with COVID-19. The interviews were conducted physically by at least two researchers and data was being digitally recorded using at least two gadgets to avoid data loss. In addition, field notes were taken later transcribed using verbatim.

### Data management and analysis

All audio-recorded data was immediately transferred after the interview from the digital recorder, and stored in a password-protected Google Drive, and later transferred to a password-protected computer. We limited access and management to the researchers only. Participants were identified by a code and not by their actual names.

Data analysis was done manually following the protocol suggested by Braun and Clarke (14). The full literal transcription of each interview, the researchers’ field notes and their descriptions were all collected to perform a qualitative analysis. Interviews were transcribed using verbatim. The following six steps of thematic analysis were undertaken; familiarization with data, generation of initial codes, searching for themes, reviewing potential themes, defining and naming themes, and writing up (producing the report). An inductive approach to coding data was used. Every principal investigator performed the initial coding of the whole data which was then discussed with the entire research team to ensure that the codes were grounded in the data. The initial codes were descriptive and provided the summary of each portion of the data. The descriptive codes which had similar or related meanings were then grouped into interpretative or latent codes. These latent codes identified the meanings that lay beneath the descriptive codes linking them together. Themes were then constructed from the descriptive and interpretative codes in an iterative process. Coded data was reviewed for similarity and overlap. Codes which clustered around a similar issue were grouped together in one theme. The relationship between themes and how they combine to produce an overall narrative were explored. The initial themes were reviewed by the study team to ensure that they reflected the original data. Some themes were subsequently left as they were, others collapsed together or split depending on their coherence and underlying meaning. The resulting themes were then defined, named, and made specifically by highlighting the unique meaning of each in line with the research objectives. Finally, the narrative report was produced with nuanced illustrations.

### Rigor of findings

Trustworthiness or rigor to ensure the quality of a study was achieved by adopting the criteria outlined by Lincoln and Guba (15), which are accepted by many qualitative researchers. These criteria include credibility, dependability, confirmability, transferability and authenticity.

Credibility was achieved through prolonged engagement with participants, where researchers spent extensive time with participants (minimum of 1 hour 30 minutes). This prolonged engagement ensured that the data collected was rich and in-depth. In addition, Triangulation was also used, where the researchers used multiple data sources, including interviews, observations and field notes and documentation of the findings. Triangulation ensured that the findings were consistent across different data sources which in turn ensured credibility of the findings. Saturation of the data also contributed to the credibility of the results, which ensured that the data was comprehensive and represented the experiences of physiotherapists.

To attain transferability, the researchers clearly defined the methodology, data collection process, including the details of participants, setting, and data collection methods that were used in this research. This clarity enables other researchers to understand the context and apply the findings to similar settings. In addition, by using purposive sampling method, the researchers ensured that the sample was relevant and informative for the research question.

Confirmability was achieved through maintenance of methodology throughout the research process, ensuring that the data collection and analysis processes were systematic and unbiased. In addition, thick description of data, and reporting of any changes that were taking place during data collection as well as in the methodology ensured that everything is available to enable other researchers to understand the context and verify the findings.

Bracketing also contributed to the confirmability of the findings whereby researchers acknowledged and set aside personal biases and assumptions, ensuring that the data analysis was objective and unbiased. Furthermore, induction analysis of data was used just to leave out researchers’ thoughts outside the data that was explored and this contributed to unbiased findings of the research.

Dependability was achieved through well-presented research methods which helped the researchers to clearly describe the research methods, including data collection and analysis procedures, enabling other researchers to evaluate and replicate the study to another setting. Pilot testing of the study also helped to accomplish dependability of this study because it ensured that the data collection tools and methods were effective and reliable. Furthermore, data saturation also contributed to dependability of this study, because the researchers continued data collection until saturation was reached, to ensure that data was comprehensive and reliable.

## Results

A total of 11 physiotherapists participated in the study. Nine participants were recruited from Queen Elizabeth Central Hospital (QECH), while two were recruited from Zomba Central Hospital (ZCH). Most participants were male (n=9) with only 2 females. Their ages ranged from 28 to 42 years, with most in their 30s. All participants were physiotherapists by occupation, with at least four years of work experience.

Participants shared diverse experiences related to their involvement in managing patients with COVID-19 at Queen Elizabeth Central Hospital and Zomba Central Hospital. These experiences have been analyzed and categorized into six main themes and thirteen sub-themes. The themes are grouped into positive and negative aspects, as well as recommendations for future improvements in the management of similar healthcare crises.

The positive themes highlight the significant impact of physiotherapy on patient outcomes, such as quick recovery and discharge, and the growing recognition of the physiotherapy profession by other healthcare workers. The negative themes address the challenges physiotherapists encountered, including difficulties in asserting their roles within multidisciplinary teams, inadequate communication among healthcare professionals, gaps in training specific to COVID-19 management, and insufficiencies within the healthcare system. These insufficiencies include lack of recognition for the physiotherapy profession, insufficient tools and personal protective equipment, inadequate staffing, and the absence of specific protocols or guidelines. Lastly, the participants proposed recommendations under three areas: the need for training tailored to the physiotherapy profession, improved multidisciplinary team collaboration, and strengthening the healthcare system’s preparedness and resource allocation to support rehabilitation professionals.

### Theme 1: Positive Experiences

#### (a) Positive impact of physiotherapy

The findings from the interviews illustrate the essential role physiotherapy played in managing COVID-19 patients. Participants described how physiotherapy facilitated faster recovery, improved patient outcomes, and supported hospital operations during the pandemic. Physiotherapists provided vital rehabilitation interventions, focusing on restoring patients’ physical functionality, respiratory strength, and overall health. These efforts were pivotal in saving lives and addressing the overwhelming demands on healthcare systems.

##### 1. Quick recovery and discharge

The study revealed that physiotherapy significantly contributed to the rapid recovery of COVID-19 patients. Through targeted interventions such as respiratory exercises, mobility restoration, and therapeutic positioning, physiotherapists enhanced patients’ recovery, enabling them to regain strength and functionality faster. This rapid improvement not only benefited patients but also helped to address the overwhelming strain on hospital resources by creating bed space for incoming patients.

> *“Patients who were receiving physiotherapy intervention were getting better much faster… Rehabilitation interventions were helping patients recover quickly, which was very good because that time there were many patients, and the space was limited.” (Participant 8, male physiotherapist, ZCH).*

These accounts emphasize how physiotherapy was critical in reducing hospital stay durations and ensuring patients regained functional independence promptly. The ability to free up hospital beds was particularly important during the pandemic, when hospitals faced capacity challenges due to the surge in COVID-19 cases. Physiotherapists’ contributions helped alleviate the pressure on healthcare facilities and allowed for better management of hospital resources.

Quick Recovery highlights the dual role of physiotherapy in improving individual patient outcomes and optimizing healthcare system efficiency. By accelerating patient recovery, physiotherapy not only enhanced the quality of care but also played a significant role in hospital operations during a time of crisis. This underscores the indispensable role of physiotherapists in managing respiratory conditions and physical deconditioning, which were common challenges among COVID-19 patients.

##### 2. Lifesaving

The findings highlight the pivotal role physiotherapists played in saving lives during the COVID-19 pandemic, particularly during critical situations where essential resources, such as oxygen, were unavailable. Participants described how physiotherapists used alternative and innovative interventions to stabilize patients’ oxygen saturation levels, preventing severe complications and potential fatalities. One of the most impactful techniques implemented by physiotherapists was prone positioning, a practice that effectively improves oxygenation in patients experiencing respiratory distress. This intervention became a lifeline during moments when oxygen supplies were depleted, showcasing the adaptability and resourcefulness of physiotherapists in crisis scenarios. One of the participants provided a striking account of this life-saving role:

> “I *remember when I was working at the isolation center at Bingu National Stadium, and all oxygen concentrators ran out of oxygen. We, physiotherapists, began putting patients in prone lying, and their oxygen saturation was improved until filled oxygen cylinders came. If this was not done, I’m sure we could have lost lives.” (Participant 7, male physiotherapist, ZCH)*.

This shows the critical role of physiotherapists in managing emergencies and adapting to resource-limited settings. Their ability to implement alternative interventions, such as prone positioning, highlights their essential contributions to the multidisciplinary management of COVID-19 patients. By leveraging their expertise, physiotherapists played a crucial role in bridging gaps in care during critical shortages, directly saving lives and preventing worse outcomes.

#### (a) Profession recognition

The study revealed that the significant contributions of physiotherapy during the pandemic led to increased recognition of the profession among other healthcare professionals and institutions. Participants noted that their role in improving patient outcomes and managing critical conditions during the pandemic helped shift perceptions and elevate the status of physiotherapy within the healthcare system.

##### 1. Recognition by other healthcare workers

Physiotherapists observed a positive shift in attitudes from their healthcare colleagues, who gained a better understanding of the role and scope of physiotherapy in patient management. By working alongside multidisciplinary teams, physiotherapists demonstrated their expertise and professionalism, earning greater respect and appreciation for their contributions.

> *“…they have now understood that physiotherapy is not just there for a show, they have understood that the role of physiotherapy is very much important because they have seen how we have demonstrated work ethics to improve the functional capacity of these patients.” (Participant 4, Male physiotherapist, QECH).*

Participants also noted that this newfound recognition extended beyond public institutions to private healthcare settings. As the impact of physiotherapy became evident, some private hospitals began referring their patients to central hospitals for physiotherapy interventions.

> *“Some patients began to be referred from Mwaiwathu private hospital to receive rehabilitation intervention here because our impact was made known to many people.” (Participant 10, Male physiotherapist, ZCH).*

The recognition of physiotherapy during the pandemic demonstrates how extraordinary circumstances can bring visibility to underappreciated professions. This increased recognition from colleagues and institutions highlights the value of physiotherapists’ expertise and their integral role in multidisciplinary teams. Furthermore, the referral of patients from private hospitals underscores how physiotherapy gained a reputation for delivering effective care, which may have long-term implications for the profession’s development and integration into healthcare systems.

### Theme 2: Negative Experiences

The COVID-19 pandemic posed significant challenges across healthcare systems worldwide. Among those deeply impacted were physiotherapists, whose roles, contributions, and needs often went unrecognized. This article explores the major challenges faced by physiotherapists as revealed in three sub-themes under the broader category of negative themes: multidisciplinary team challenges, training gaps, and healthcare system insufficiencies.

#### (a) Multidisciplinary team challenges

##### 1. Difficulty in Asserting Rehabilitation Professional Roles

Physiotherapists faced significant challenges in asserting their roles within multidisciplinary teams. Many healthcare professionals, including doctors and nurses, had limited understanding of the scope of physiotherapy practice. This lack of awareness led to misunderstandings and undervaluation of physiotherapists’ contributions.

> “*I think most people don’t fully understand what physiotherapy is all about. They do not know what it is that we do, to the extent that they are not aware of the scope of our practice.” (Participant 7, Male Physiotherapist, ZCH)*

This misunderstanding also extended to managing COVID-19 patients, as some team members questioned physiotherapists’ involvement.

> *“…Sometimes the doctors and nurses could not understand why we were supposed to go into the tents or COVID-19 camps to attend to patients with COVID-19, and they did not understand our role as physios.” (Participant 11, Female Physiotherapist, QECH)*

The undervaluation of physiotherapists stemmed from a lack of knowledge and communication about their specialized skills. This created barriers in collaborative care, leading to missed opportunities for rehabilitation interventions that could improve patient outcomes.

##### 2. Inadequate Communication

Another barrier was insufficient communication among multidisciplinary teams. Separate ward rounds by different professions restricted interdisciplinary discussions and collaboration.

> *“Health care professionals were doing ward rounds separately. For example, doctors could do their own rounds, same as rehabilitation professionals. This brought about challenges because rehabilitation professionals could not meet other healthcare professionals to discuss about some things concerning the patients or to consult.” (Participant 3, Male Physiotherapist, QECH)*

Poor communication and siloed practices undermined team dynamics and created challenges in providing cohesive patient care. Interdisciplinary collaboration is essential for managing complex cases like those seen during the pandemic.

#### (b) Training Gap

##### 1. Inadequate Training on COVID-19 Management

Physiotherapists reported that they were inadequately prepared to manage COVID-19 patients due to a lack of targeted training programs. Most training sessions were general and did not address the specific needs of rehabilitation professionals.

> *“…The trainings were not tailor-made. After the training, at least they could have isolated and said, ‘Okay, this is how you are supposed to handle a COVID-19 patient.’ They were supposed to have trainings that were tailored to rehabilitation.” (Participant 6, Male Physiotherapist, QECH)*

Many physiotherapists felt that training was too slow to adapt to the pandemic’s needs, leaving them uncertain about how to handle patients presenting with unique COVID-19 symptoms.

> *“…We were not quick enough in terms of the trainings on how we can manage patients with COVID-19… When there is a pandemic, there is a need to quickly introduce rehabilitation interventions tailored to treatment modalities for such a pandemic.” (Participant 4, Male Physiotherapist, QECH)*

The lack of specific training for physiotherapy professionals highlights systemic gaps in pandemic preparedness. Tailored training could have equipped physiotherapists with the necessary skills and confidence to handle the unique challenges of COVID-19 patients.

#### (c) Healthcare System Insufficiency

##### 1. Lack of Recognition

Physiotherapists were not included in the government’s initial recruitment of healthcare workers to manage COVID-19, reflecting the lack of recognition for their role.

> *“We went to the hospital director to ask him why rehabilitation professionals were not included among healthcare workers who were managing COVID-19 patients, and he said, ‘If it is allowance you are looking for, we can recruit you as clinicians.’” (Participant 10, Male Physiotherapist, ZCH)*

Even during public recognition events, such as when the president awarded medals to healthcare professionals, physiotherapists were excluded.

> *“…The president gave out medals to health workers. I think there was not a physiotherapist there? But regardless, they did not fully recognize us.” (Participant 7, Male Physiotherapist, ZCH)*

The lack of recognition undermined physiotherapists’ morale and limited their ability to advocate for their profession’s essential role in healthcare. Recognizing their contributions could have encouraged better resource allocation and integration into pandemic responses.

##### 2. Insufficient Tools and Personal Protective Equipment

Physiotherapists reported shortages of critical equipment including personal protective equipment (PPE) necessary for managing COVID-19 patients.

> *“When it comes to management, most of the rehabilitation equipment was not available… Spirometers were not available during the management in the wards.” (Participant 6, Male Physiotherapist, QECH)*
>
> *“…We struggled to have PPEs, and we had a strike to get personal protective equipment, so we were not prepared enough.” (Participant 4, Male Physiotherapist, QECH)*

The lack of essential resources hindered physiotherapists’ ability to provide effective care, increasing their risk of exposure and reducing patient care quality.

##### 3. Insufficient Staff

The shortage of physiotherapists exacerbated the challenges, with one physiotherapist often tasked with managing large numbers of patients.

> *“… One of the challenges is staff… There were too few physiotherapists working in the system… One physio against 40 breathless patients at one given time is too much for anybody to manage.” (Participant 7, Male Physiotherapist, ZCH)*

The lack of sufficient staff not only compromised the quality of care but also placed an immense physical and emotional burden on physiotherapists.

##### 4. Lack of Rehabilitation-Specific Protocols

Physiotherapists operated without clear guidelines or protocols tailored to COVID-19 rehabilitation, leading to inconsistent approaches.

> *“In terms of guidelines, we didn’t have any… The training we received didn’t produce guidelines to guide physiotherapists in managing COVID-19.” (Participant 5, Male Physiotherapist, QECH)*

The absence of standardized protocols created variability in patient care and left physiotherapists uncertain about best practices.

The challenges faced by physiotherapists during the COVID-19 pandemic highlight systemic gaps in healthcare systems, including inadequate recognition, insufficient training, and lack of resources. Addressing these issues requires greater collaboration, targeted training programs, sufficient staffing, and the development of rehabilitation-specific guidelines. Recognizing and supporting physiotherapists is essential for building resilient healthcare systems capable of responding to future pandemics.

### Theme 3: Participants’ Recommendations

Drawing from the experiences of physiotherapists in multidisciplinary teams, their insight into the healthcare system’s response to COVID-19, and the training they received, several recommendations were proposed to improve the management of COVID-19. These recommendations address training needs, the role of multidisciplinary teamwork, and necessary improvements in the healthcare system.

#### 1. On Training

Participants emphasized that future training for physiotherapists should be profession-specific and tailored to the needs of COVID-19 patients.

> *“The training we received was general… I suggest training should focus on rehabilitation professionals specifically…”(Participant 11, Female Physiotherapist, QECH)*
>
> *“…. We need training that is specific to physiotherapy… we need to know how to manage patients with COVID-19, specifically…”(Participant 9, Male Physiotherapist, QECH)*

#### 2. On Multidisciplinary Teamwork

Participants highlighted the importance of regular multidisciplinary team meetings to share knowledge and skills, fostering better collaboration and understanding among team members.

> *“…I recommend multidisciplinary team meetings to share skills and knowledge on managing conditions…”(Participant 8, Male Physiotherapist, QECH)*
>
> *“…We need to work together as a team… we need to share our knowledge and skills to provide better care for patients with COVID-19…”(Participant 10, Male Physiotherapist, ZCH)*

#### 3. On the Healthcare System

Participants stressed the need for the healthcare system to be consistently prepared for health emergencies by ensuring sufficient resources and facilities. They also recommended increasing the recruitment of rehabilitation professionals to meet the demands during crises like COVID-19.

> *“…The healthcare system should be always prepared for health emergencies in terms of resources and facilities…”(Participant 3, Male Physiotherapist, QECH)*
>
> *“…The government could have improved recruitment of healthcare personnel, especially rehabilitation professionals, who were overwhelmed by the increased number of patients with COVID-19…”(Participant 6, Male Physiotherapist, QECH)*
>
> *“…We need more physiotherapists… we need more rehabilitation professionals to manage patients with COVID-19…” (Participant 9, Male Physiotherapist, QECH)*

## Discussion

This study aimed to explore the experiences of physiotherapists managing COVID-19 patients in central hospitals in Malawi, uncovering both challenges and successes. The findings revealed a range of experiences, including multidisciplinary challenges, healthcare system insufficiencies, training gaps, and positive impacts of rehabilitation interventions. These results align with and differ from other studies conducted globally.

The difficulty in asserting physiotherapy roles was a recurring theme in this study, as physiotherapists faced limited recognition from other healthcare professionals. This finding is consistent with a Nigerian study by Chinoso et al, which reported similar struggles due to the lack of understanding of physiotherapy’s scope in managing COVID-19 patients (16). However, studies in developed countries, such as the Netherlands highlighted better interdisciplinary collaboration (17), suggesting that differences in recognition stem from the limited awareness and integration of physiotherapy in healthcare systems of developing countries like Malawi. Inadequate communication within multidisciplinary teams emerged as a significant challenge, driven by the need to avoid ward congestion during the pandemic. This fragmentation limited physiotherapists’ opportunities for referrals, leading to missed rehabilitation opportunities. In contrast, the study conducted in Netherlands reported effective communication and teamwork, underpinned by established protocols and better organizational structures, which facilitated smoother collaboration (17). This disparity reflects the lack of robust systems in Malawi’s healthcare framework to promote coordinated care during emergencies.

Healthcare system insufficiencies were evident, including a lack of recognition, insufficient tools, and inadequate staffing. The lack of physiotherapists in frontline roles during the initial wave mirrored findings in Nigeria, where rehabilitation professionals were overlooked and had to advocate for their inclusion (16,18). Similarly, shortages of personal protective equipment (PPE) in Malawi reflected global trends, as seen in a U.S. study by Campo et al. (19), although developed countries were generally better equipped by the second wave. The insufficiency of staff, particularly physiotherapists, in Malawi aligns with local studies highlighting the chronic understaffing of healthcare workers, exacerbated by the pandemic (20)(21). This contrasts with developed countries, where the workforce was sufficient despite higher COVID-19 case numbers (19), showcasing pre-existing disparities in healthcare infrastructure.

The lack of rehabilitation-specific protocols during the pandemic forced physiotherapists to develop center-based guidelines. This inconsistency contrasts with findings from the Netherlands, where well-organized rehabilitation protocols were in place (17), reflecting the advanced state of physiotherapy in developed countries. This highlights the need for Malawi to establish standardized protocols to ensure consistent care. Inadequate training specific to COVID-19 management hindered physiotherapists in Malawi. Although they possessed general respiratory care knowledge, the unique challenges of COVID-19 required tailored training, which was lacking. This gap is a common issue in low-income countries, whereas developed countries provided more targeted training due to their advanced health systems and resources (19)(17).

Despite these challenges, the study revealed the positive impacts of physiotherapy, such as quick patient discharges and lifesaving interventions like prone positioning and chest physiotherapy. These findings align with existing literature emphasizing the critical role of physiotherapy in enhancing recovery and reducing mortality in COVID-19 patients (22).

In summary, our findings align with global studies in highlighting the challenges faced by physiotherapists, though the degree of difficulty is exacerbated in Malawi due to systemic gaps. The lack of recognition, inadequate communication, and insufficient resources reflect the broader challenges faced by developing countries, while the positive outcomes of physiotherapy interventions underscore their indispensable role in healthcare systems worldwide. Addressing these issues requires better integration of physiotherapy into healthcare systems, improved training, and enhanced resources to optimize care during future health emergencies.

### Study strengths and limitations

The use of in-depth interviews for data collection in this qualitative study (rather than questionnaires used in quantitative studies) ensured collection of comprehensive data from the participants. This ensured researchers’ comprehensive understanding of the phenomenon under study. The study was also conducted at two tertial referral hospitals in the southern region of Malawi, where all cases of COVID-19 in the region of Malawi were being referred to, and these hospitals are the centers where expert care of COVID-19 patients were being offered. Participants’ experiences may, therefore, be representative of other physiotherapists’ experiences in other regions of the country. Furthermore, the use of pilot-tested topic guide which ensured the credibility of the interview guide before using it on actual participants. Moreover, data saturation was reached, which ensured the investigators that enough data from the participants had been collected.

However, participation in this study was dominated by the male gender as they were the ones who gave consent in larger numbers than female gender. This limits the generalizability of the predominantly male physiotherapists’ experiences to female physiotherapists. The generalizability of the findings to all physiotherapists across the country is also limited by the fact that data were collected from physiotherapists working at central hospitals in the southern region only. Significantly, the study took place two years post-pandemic. This timing may have introduced recall bias, potentially compromising participant data accuracy due to diminished long-term memory retrieval.

### Conclusions

This study explored the experiences of physiotherapists managing COVID-19 patients in Malawi. Findings show that the pandemic positively increased recognition of physiotherapy in respiratory rehabilitation and critical care and contributed to improved patient recovery outcomes. However, participants also reported significant challenges including limited recognition within multidisciplinary teams, inadequate COVID-19-specific training, and shortages of equipment (including PPE) and staffing. To strengthen pandemic preparedness, the study recommends that Malawi’s healthcare system enhances interdisciplinary collaboration, provides targeted training for rehabilitation professionals, and ensures adequate resources and physiotherapy-specific treatment protocols. Addressing these gaps will improve the integration and delivery of rehabilitation services and support better long-term patient outcomes.

## Data Availability

All data generated and analysed during this study are included in this article. Its supplementary information files (including data collection tools and consent forms) are available from the corresponding author on reasonable request.

